# A Cross-Sectional Study of Parenting Styles Among Working Mothers: Trends and Determinants

**DOI:** 10.1101/2025.04.05.25325259

**Authors:** Muhammad Shaheryar Bashir, Moniba Iqbal, Talha Mahmood, Abdullah Khalil

## Abstract

**Introduction:** The development of children is greatly influenced by the parenting styles employed; authoritative, authoritarian, and permissive approaches all have different effects on behavior and self-esteem. This study fills a knowledge gap in Rawalpindi, Pakistan by identifying different parenting styles of working mothers in Rawalpindi.

**Objectives:** The objectives of this study are to determine how often working women in the Rawalpindi district choose a particular parenting style, to measure the association between type of parenting style and profession, to check the effect of working hours on adoption of parenting style by working women.

**Operational Definitions:** *Parenting styles:* Parenting style refers to the approach or manner in which parents interact with their children and make decisions regarding their upbringing. This includes aspects such as disciplinary strategies, communication patterns, and levels of warmth and support.

*Working women:* Working women are those who engage in employment outside the home, they contribute in the workforce and economy in various professions and have working hours ranging from 6-8 hours/day.

**Materials and Methods:** This three-month Cross-sectional study in Rawalpindi was specifically focused on working women. 221 participants who agreed to participate were working mothers with children who fulfilled the inclusion criteria using a sample size of 289 and a questionnaire-based survey. The study eliminated women who failed to provide informed consent or who gave insufficient data. SPSS analysis demonstrated proportion of different parenting styles among Rawalpindi’s working mother and the association with various demographic factors.

**Results:** Of the 221 working women in Rawalpindi who responded, 86.45% had an authoritative parenting style, 11.3% had an authoritarian one, and 2.3% had a permissive one. There was a substantial association found between authoritative parenting and living in an urban area, earning more money, becoming a teacher, and having fewer kids. Participants who had divorced were more inclined to be lenient. Parenting methods did not significantly change according to working hours or educational attainment.

**Conclusions:** The results of this study show that the authoritative parenting style is the most common among working mothers in Rawalpindi. A significant link was found between the number of children, marital status, and family income. Promoting a healthy home environment and better cognitive and mental development for children, stakeholders must create educational programs, counselling services, and recreational facilities in order to promote good parenting.

## INTRODUCTION

Parenting styles are strategies used by parents to raise their children, playing a significant role in developing a child’s personality and shaping their social skills. Baum rind’s classification categorizes parenting styles into authoritarian, authoritative, and permissive styles, each with unique influences on children’s demeanor and self-esteem.

Parents adopt different parenting styles based on factors such as financial status, stress, and cultural background. Studies have shown that authoritative parenting is dominant among younger parents in Saudi Arabia, with family size and husbands’ income being the only significant factors. Children with authoritative parents exhibit more positive behavior and are more likely to attend daycare.

Working mothers tend to adopt authoritative parenting styles, with moderate levels of parenting satisfaction reported. Children with parents, who adopt authoritative parenting style, tend to have a higher self-esteem thus establishing a positive correlation between this parenting approach and self-esteem in children.

Parenting is a continuous and dynamic process, with actions impacting multiple facets of a child’s development from birth to adulthood. In recent years, urbanization has led to a decrease in indulgence in household activities and quality child care, which is said to impact parents’ parenting style. However, there is a significant research gap regarding this topic in Pakistan, particularly in the Rawalpindi district.

## METHODS

This study is designed as a descriptive cross-sectional study, set within the district of Rawalpindi, and will span approximately three months following approval from the Ethical Review Board. The study population comprises working women from various sections of Rawalpindi, aiming to include a sample size of 285 participants. This number is calculated using the WHO calculator, ensuring a 95% confidence interval and a 5% margin of error. The sample size is calculated using 75% as anticipated frequency from our reference study, the one done in Saudi Arabia ^[15]^ The sampling technique employed is non-random convenience sampling. Participants will be selected based on their availability and consent. Consent will be obtained through a consent form attached to the questionnaire. Data will be collected using a structured questionnaire developed by the researchers, which will serve as the primary data collection tool.

The inclusion criteria for the study are as follows: working parenting women residing in Rawalpindi, women with an education level of matriculation or higher, women working a minimum of 6-8 hours per day, and women with children studying in classes 1 to 10. The exclusion criteria are: women who work from home, women who have hired caretakers, working women whose children live in boarding schools, working women whose children live with their grandparents, and women with mentally retarded or handicapped children.

The tests applied in our research included T-test, ANOVA, MANN WHITNEY U TEST, KRUSKAL WALLIS TEST.

## Results

Of the 246 responses received, 25 were found to be incomplete and consequently excluded. The most common parenting style in our study population was authoritative (191, 86.4%), followed by authoritarian (25, 11.3%), while only 5 (2.3%) used the permissive style (Table 1). (Figure 1).

**Table 1.** Frequency of parenting style

| Type | Frequency | Percentage |
| --- | --- | --- |
| Authoritative | 191 | 86.4% |
| Authoritarian | 25 | 11.3% |
| Permissive | 5 | 2.3% |

**FIGURE 1.**
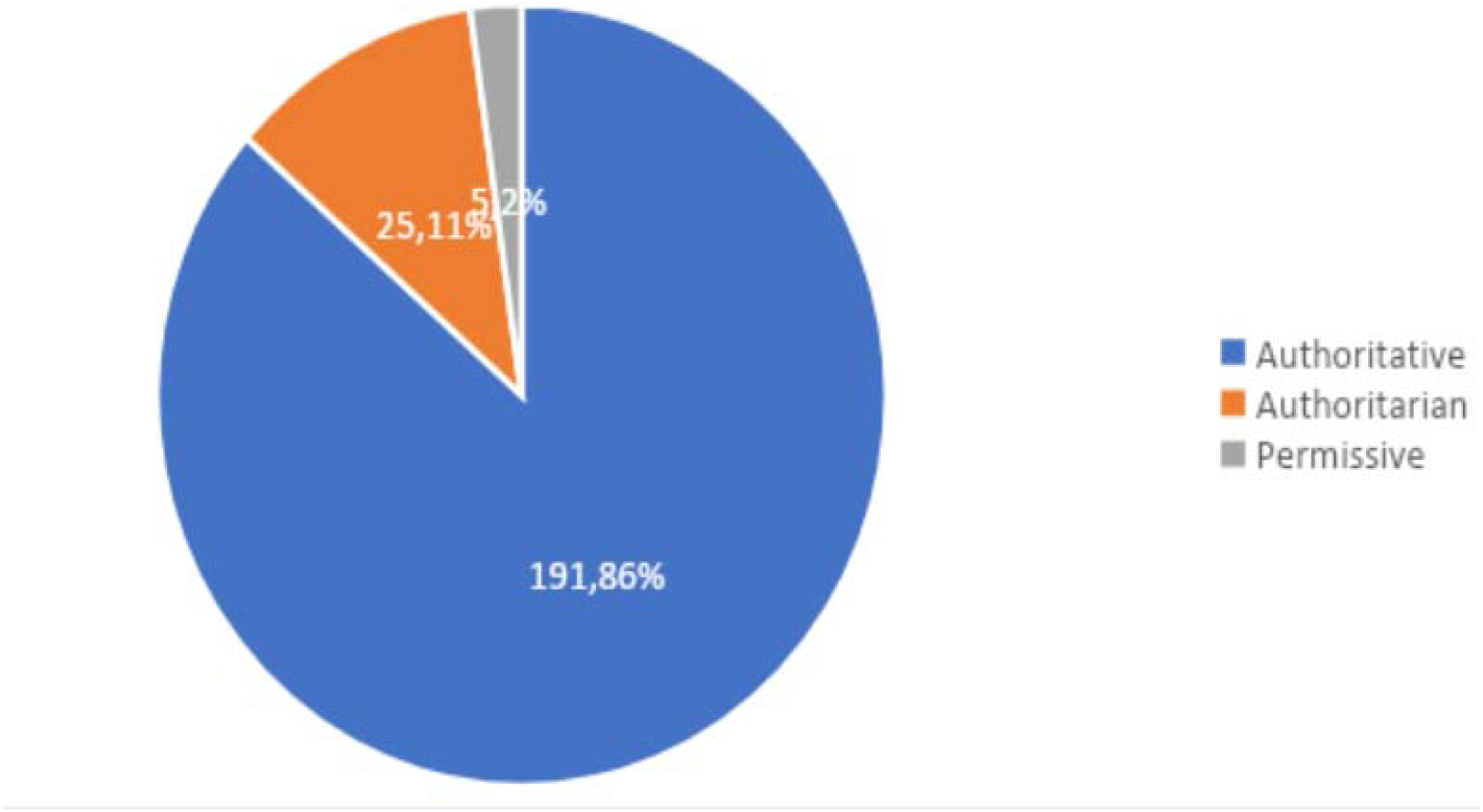
Frequency of parenting styles based on Baumrind’s classification (n=221)

The AUTHORITARIAN parenting style data was normal and ANOVA test was applied. Wherea AUTHORITATIVE parenting style data was negatively skewed and PERMISSIVE parenting style data was positively skewed and for these two parenting styles KRUSKAL WALLIS test was applied

**Table 2.** Participants’ demographic characteristics (n=221)

| **Item** | **Number** | **Percentage** |
| --- | --- | --- |
| Residence |  |  |
| Rural | 24 | 10.9 |
| Urban | 197 | 89.1 |
| Marital status |  |  |
| Married | 187 | 84.6 |
| Divorced | 22 | 10 |
| Widow | 12 | 5.4 |
| Level of Education |  |  |
| Intermediate or below | 6 | 2.7 |
| Bachelors | 55 | 24.9 |
| Masters | 160 | 72.4 |
| Occupation |  |  |
| Teaching profession | 155 | 70.1 |
| Medical profession | 42 | 19 |
| Others | 24 | 10.9 |
| Family income |  |  |
| Below Rs.50,000 | 36 | 16.3 |
| Rs.50,000-Rs.80,000 | 57 | 25.8 |
| Rs.80,000-Rs.100,000 | 70 | 31.7 |
| Above Rs.100,000 | 58 | 26.2 |
| Number of children |  |  |
| ≤3 | 161 | 72.9 |
| >3 | 60 | 27.1 |
| Work hours |  |  |
| ≤6 hours | 111 | 50.2 |
| >6 hours | 110 | 49.8 |

**FIGURE 2.**
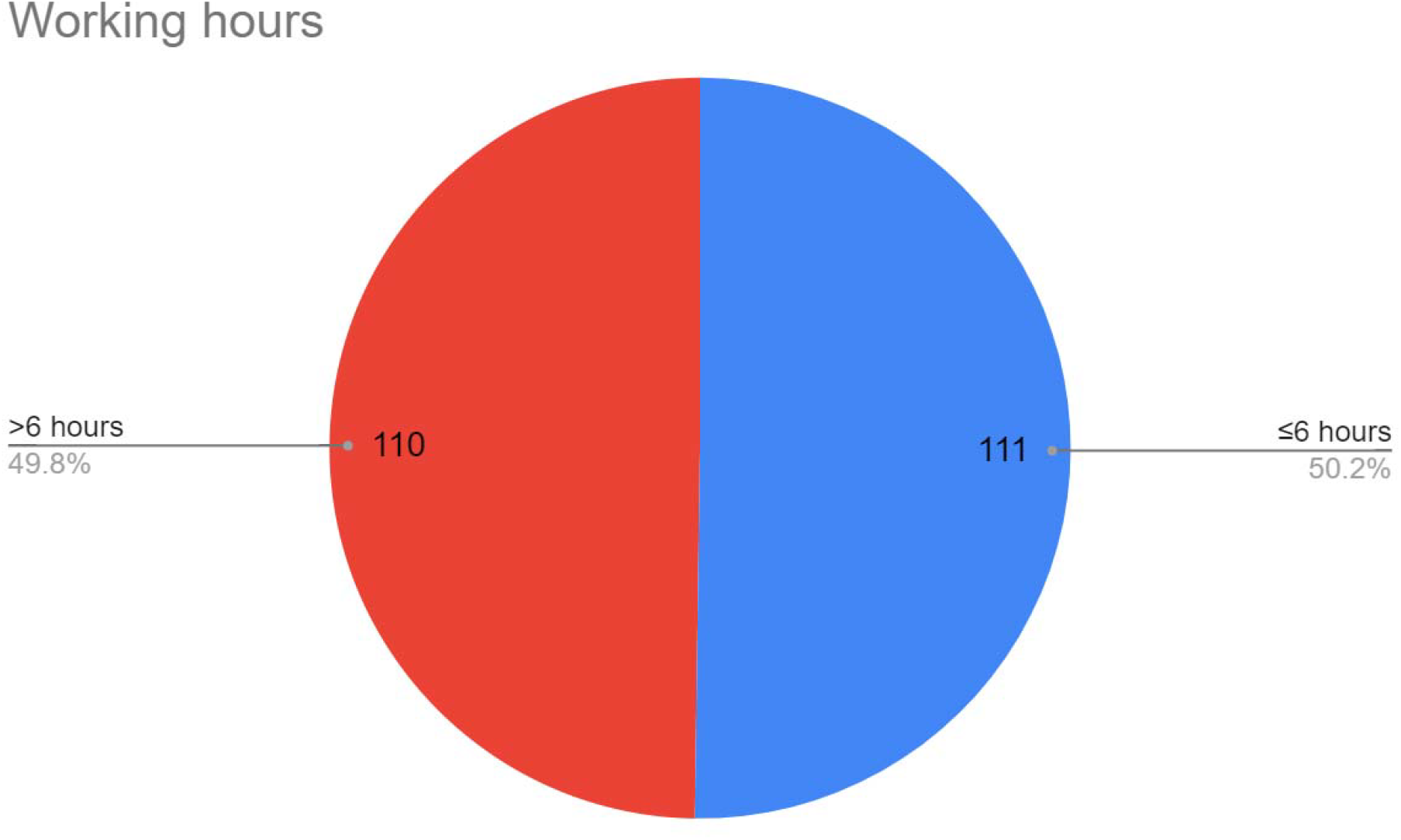
Working hours of most of the women were less than six hours (111, 50.2%) while another half had more than six hours (110, 49.8%).

**FIGURE 3.**
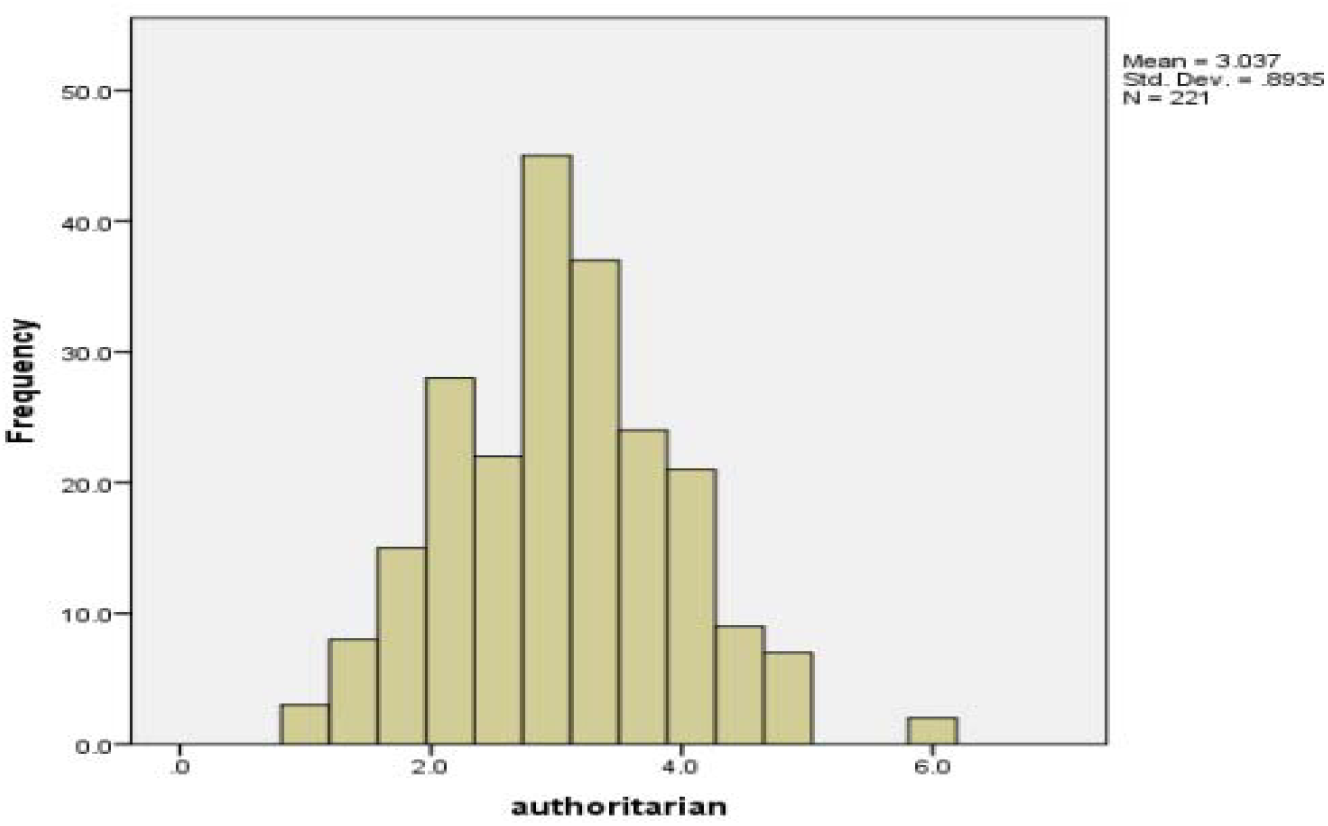
Frequency distribution for authoritative and permissive style was negatively and positively skewed respectively while it followed normal distribution in case of authoritarian parenting style Table 3 shows the comparison between the median and interquartile range of each parenting style in terms of participants’ demographics. Mean and standard deviation was used for comparison between authoritarian and socio-demographic variables only. There were no statistical differences in parenting styles with respect to the level of education or working hours.

**Table 3.**
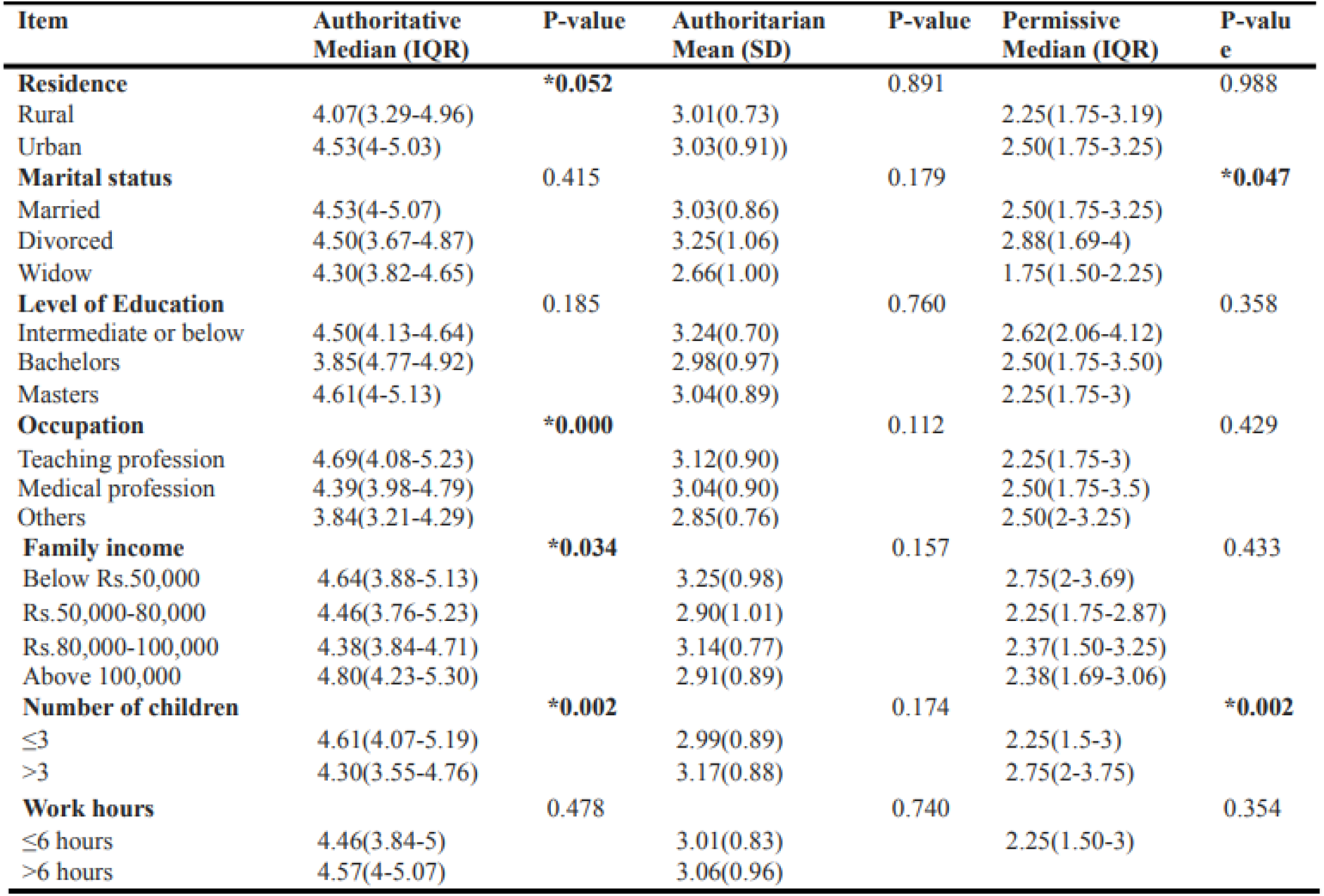
Association between participants’ sociodemographic characteristics and parenting styles (n=221)

**FIGURE 4.**
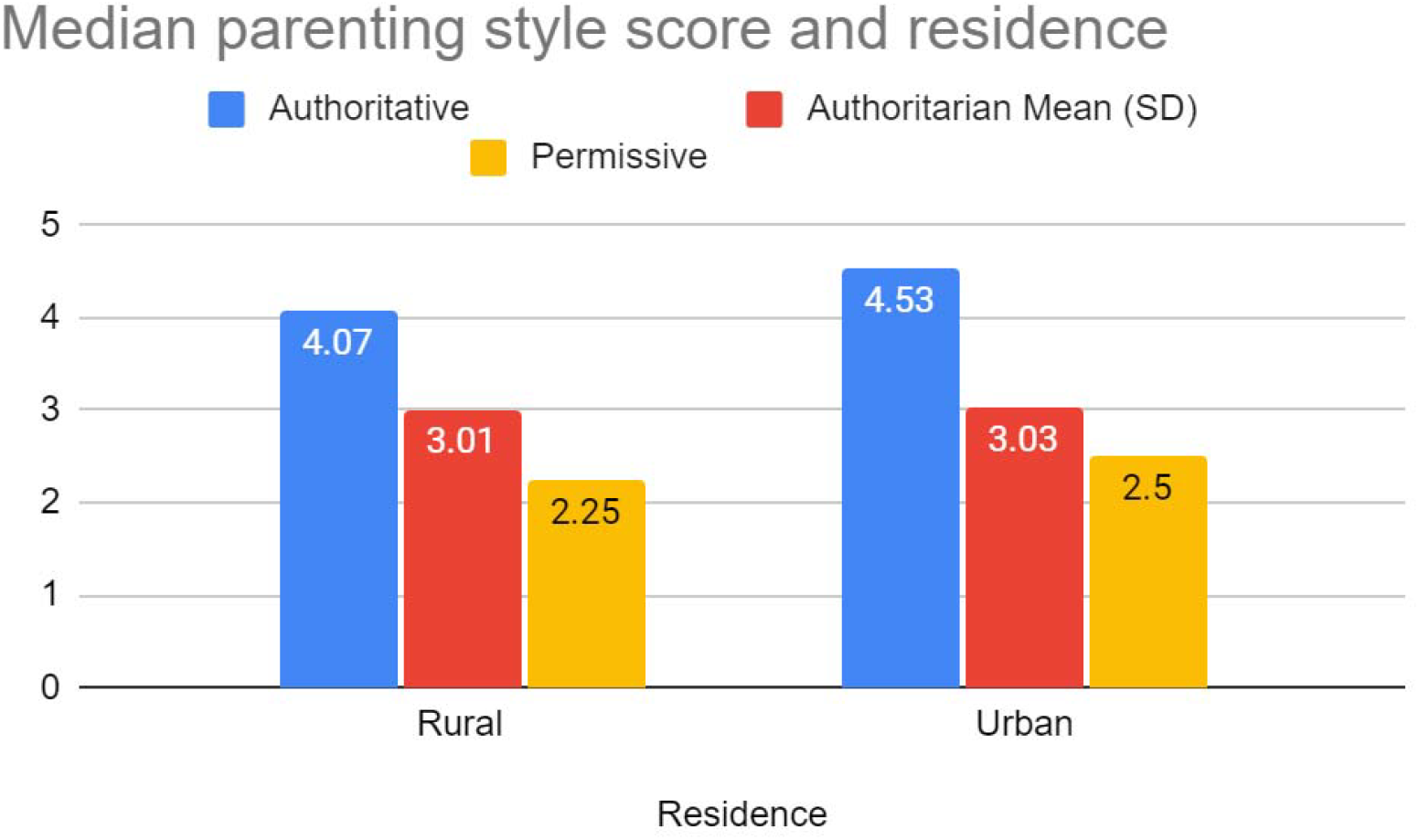
Urban residence was significantly associated with authoritative style (p=0.052), with a median score of 4.53 and an IQR of 4-5.03.

**FIGURE 5.**
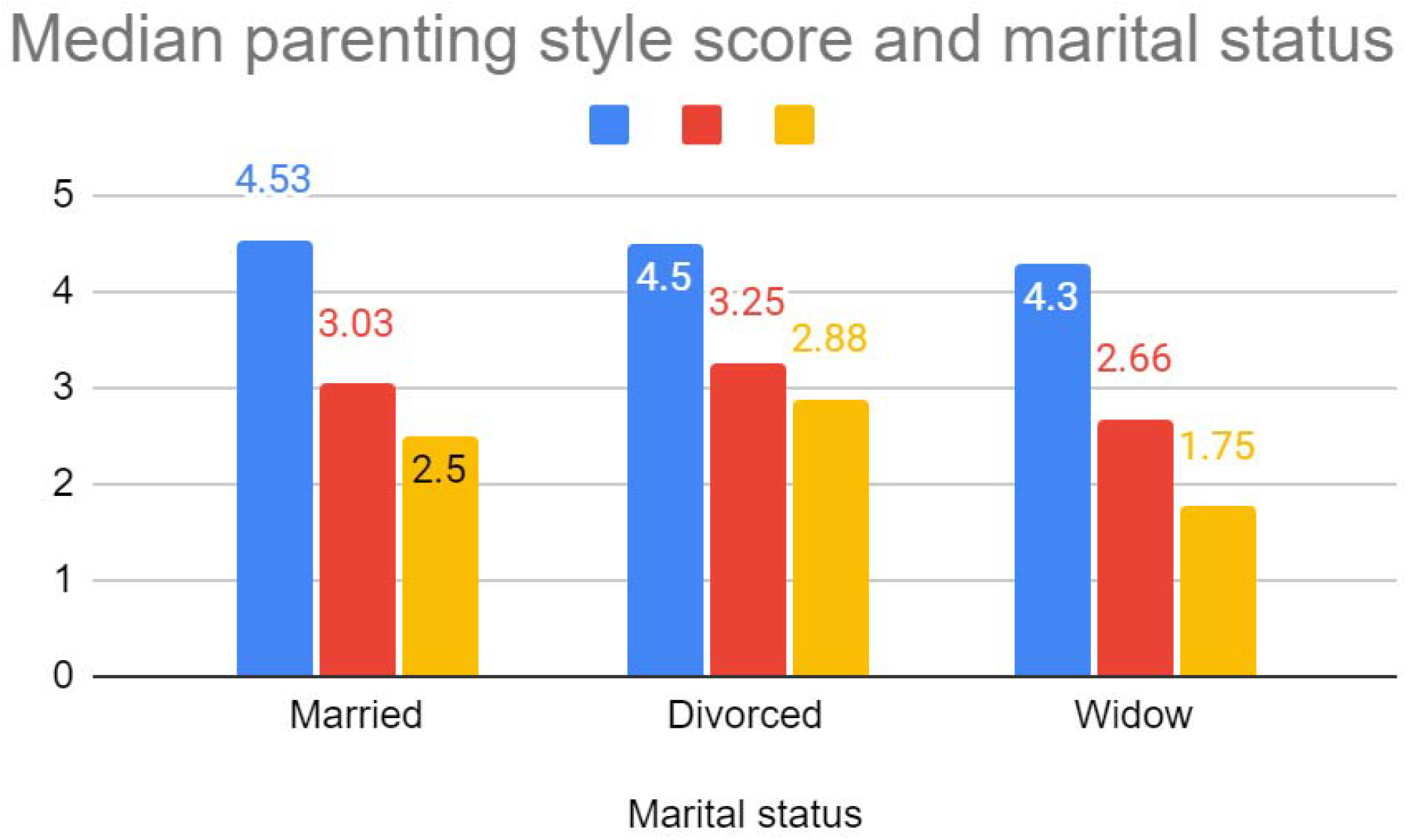
Permissive parenting was more common among divorced women, while widows were least likely to adopt a permissive style (p=0.047).

**FIGURE 6.**
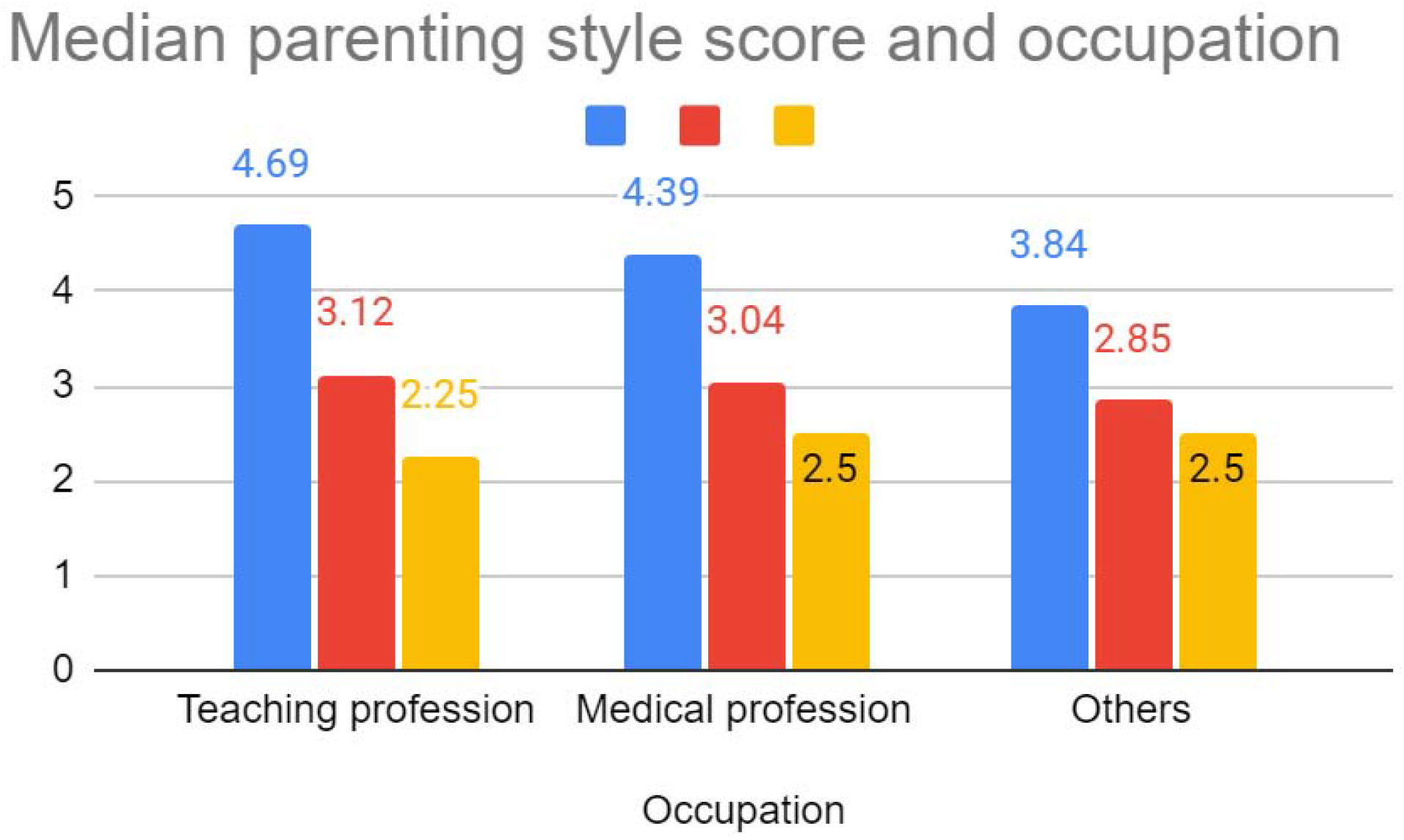
Application of Kruskal-Wallis test shows that the distribution of authoritative style is more common across the teaching profession (p=0.000).

**FIGURE 7.**
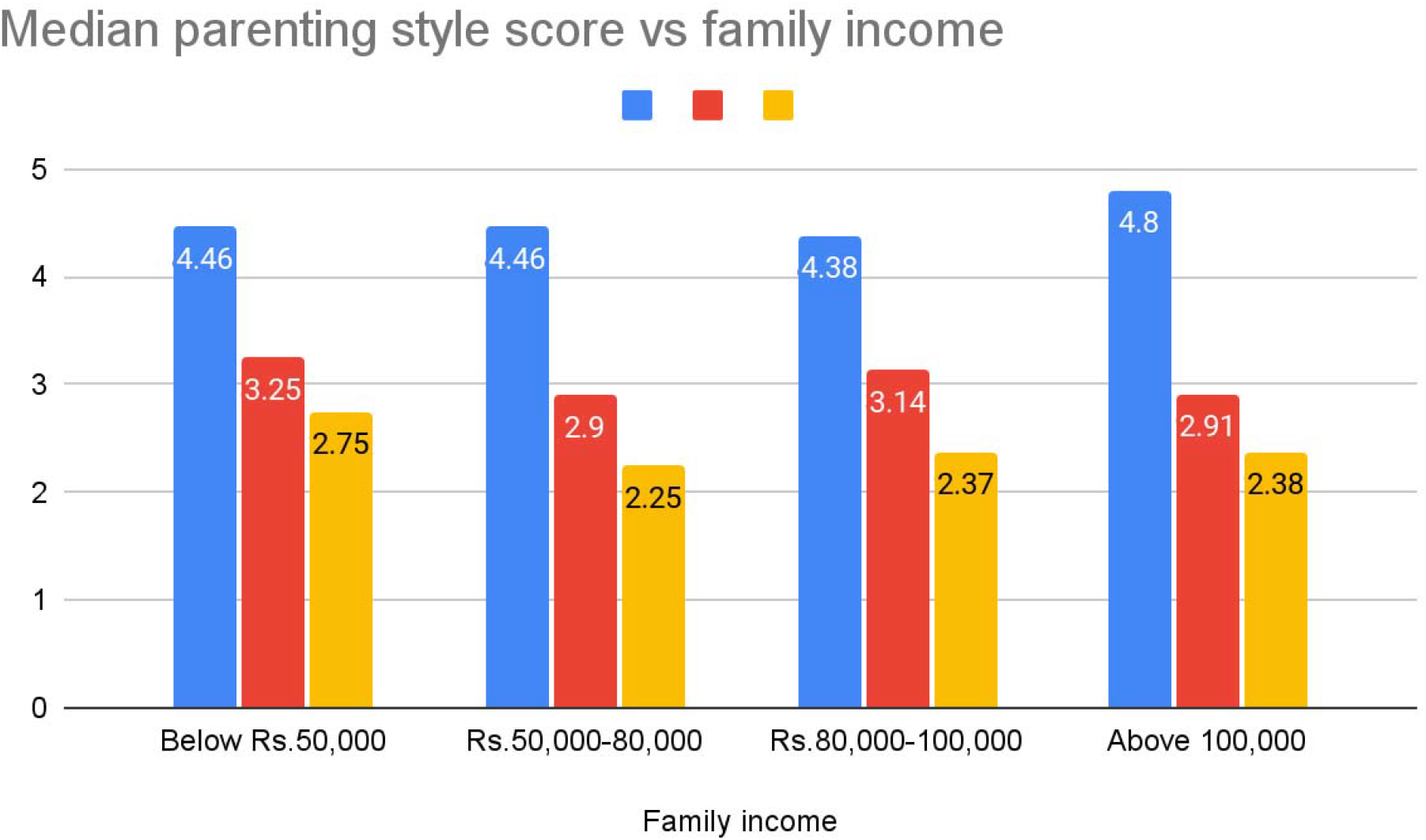
Family income had a significant association with parenting style. A family income of Rs. 100,000 or above was associated with authoritative style (p=0.034).

**FIGURE 8.**
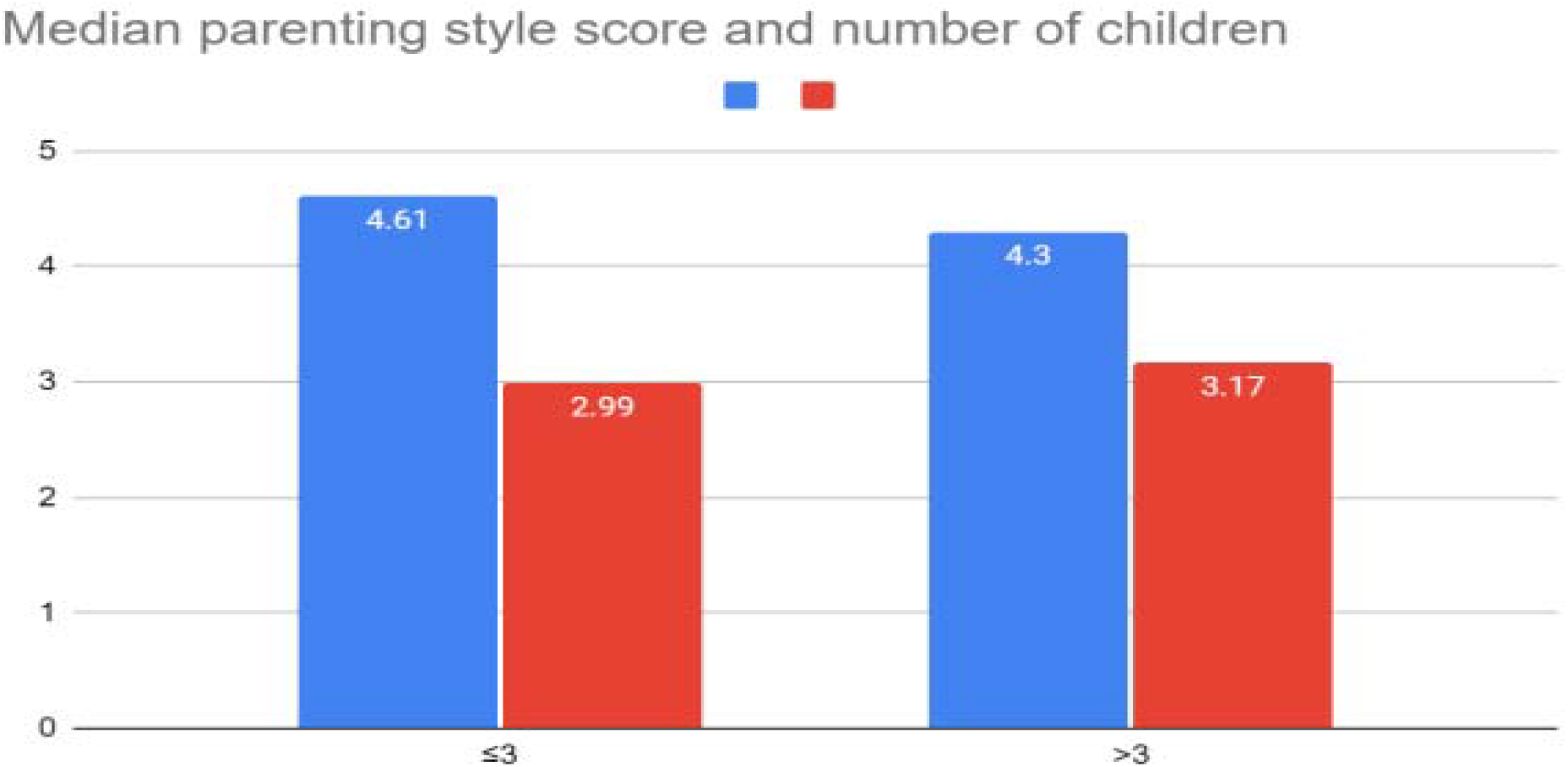
Some mothers with less than three children had authoritative parenting style while some had permissive parenting styles (p=0.002)

**FIGURE 9.**
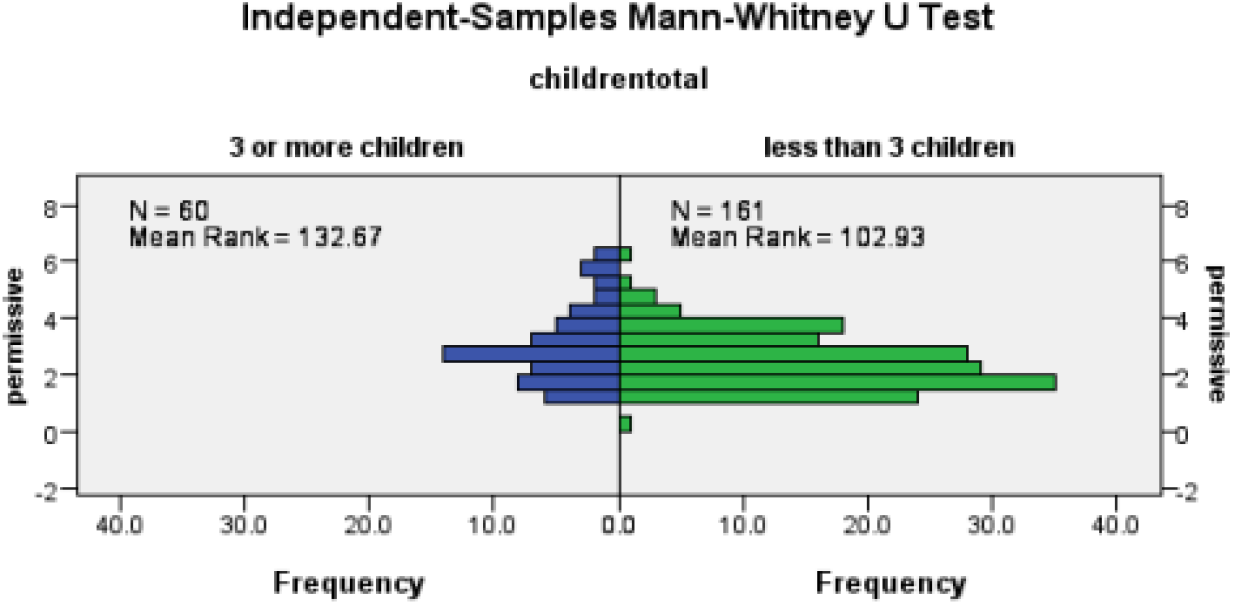
Results show significant relationship between number of children and autgoritative as well as pwrmissive parenting. Overall the authoritative style scores were high but a sifnificant association was also found between some women having more than three children and permissive parenting style. Test of correlation shows a significant moderate correlation (i.e. 0.5) between authoritarian and permissive parenting style (p=0.000) which indicates that the mothers with the authoritarian parenting style also tend to have permissive parenting style.

**FIGURE 10.**
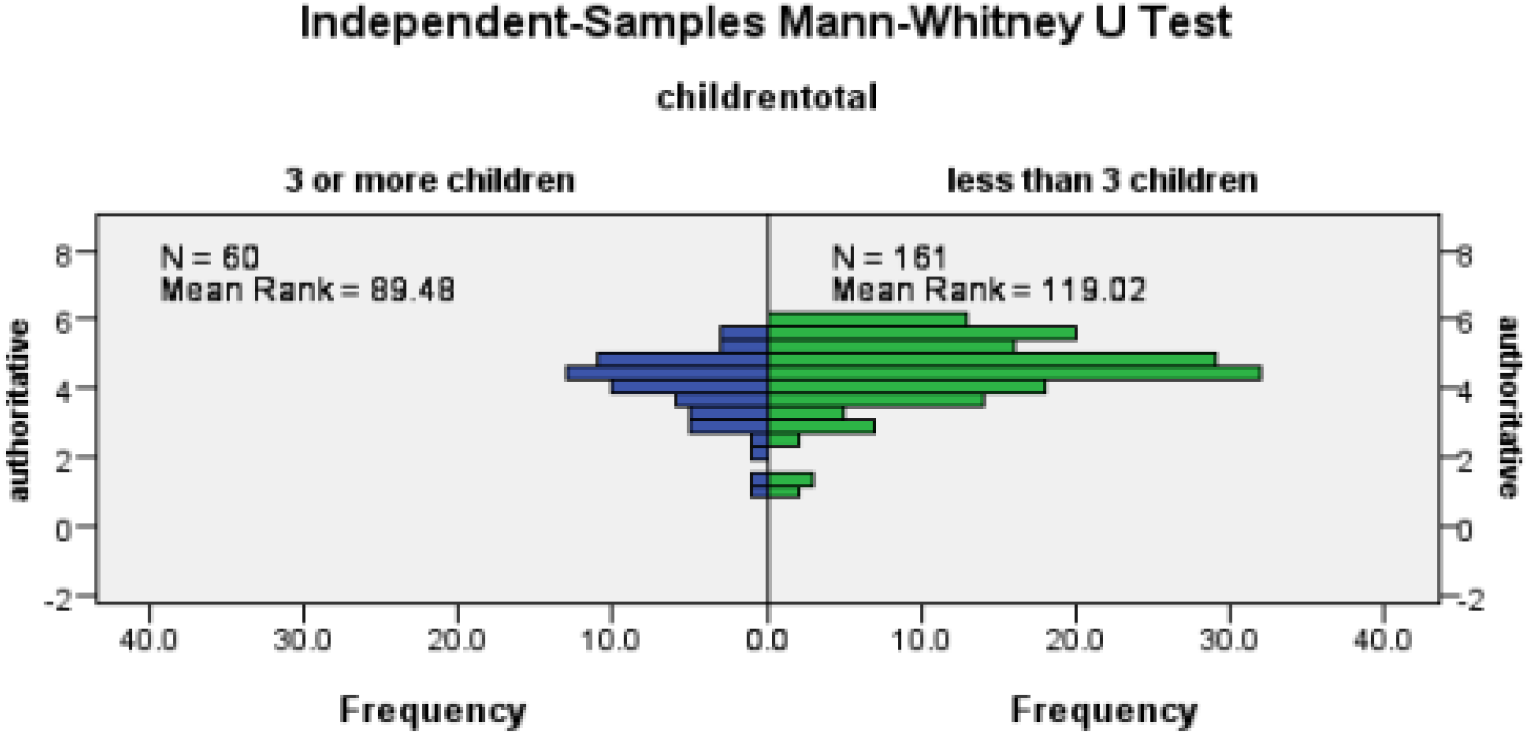

## Discussion

Parenting is a cornerstone in shaping an individual’s development and their capacity to adapt, thrive, and contribute within society. Baumrind’s seminal classification into authoritarian, authoritative, and permissive parenting styles offers a lens through which to view the complex interplay between parental behavior and child outcomes. This study highlights the predominance of authoritative parenting among working mothers in Rawalpindi, revealing significant sociocultural and demographic influences on parenting choices.

The predominance of authoritative parenting (86.4%) among working mothers can be attributed to its balanced approach, combining warmth, structure, and consistent discipline. Authoritative parents promote open communication and nurture independence while maintaining clear boundaries, which aligns with positive outcomes in self-esteem, emotional regulation, and academic performance in children, as supported by Howenstein et al. (2015).^1^ The findings resonate with studies from Arab societies, where shared cultural values emphasize the importance of structure and guidance.^2^

Urban living significantly influences parenting styles, with urban mothers showing a stronger inclination towards authoritative parenting. This association may stem from exposure to diverse parenting philosophies, improved access to educational resources, and socioeconomic stability, as noted by Martínez et al. (2007).^3^ Urban environments often provide opportunities for mothers to engage with other parents and learn effective practices, fostering an authoritative approach. In contrast, rural mothers may face limitations in resources and educational opportunities, potentially impacting their parenting strategies.^4^

Family income plays a decisive role, as higher earnings enable families to invest in enriching activities and stable home environments conducive to authoritative parenting. Almudhee et al. (2023) observed similar correlations in Saudi Arabia, emphasizing the link between financial security and proactive parenting strategies.^5^ Mothers earning above Rs. 100,000 showed a significant preference for authoritative parenting, suggesting that financial stability enables greater focus on fostering children’s development.

Marital status emerged as a crucial determinant of parenting styles. Divorced mothers showed a higher tendency towards permissive parenting, potentially reflecting efforts to compensate for disruptions in familial cohesion. This aligns with findings by Sanvictores & Mendez (2022) and Arafat & Khan (2022), which highlight the psychological factors influencing single parents, including guilt and fear of estrangement, leading to leniency in discipline.^6,7^ The emotional challenges faced by divorced parents often manifest in permissive approaches to maintain positive relationships with children.

Interestingly, the study did not find a significant association between educational attainment and parenting styles. While higher education is often presumed to enhance parenting practices, the findings reflect broader cultural dynamics that may outweigh educational influence. For example, Asian parents, regardless of educational background, often adopt authoritarian styles due to societal emphasis on discipline and conformity.^2^ In contrast, permissive parenting is more prevalent in Western contexts, driven by cultural values of individuality and autonomy.^6^

Furthermore, family size significantly impacts parenting styles. Mothers with fewer children displayed authoritative tendencies, as fewer dependents allow greater attention to individual needs. Conversely, permissive parenting was observed among mothers with larger families, possibly due to the challenges of managing multiple children simultaneously.^8^ Resource allocation and time management remain critical factors influencing parental approaches.

The findings also echo global patterns observed in similar studies. For instance, in Indonesia, authoritative parenting was associated with greater socioeconomic stability, while permissive styles were linked to familial stress and limited resources.^9^ In the USA, cultural emphasis on individuality and leniency has led to the dominance of permissive parenting, highlighting contrasting global dynamics.^6^

## LIMITATIONS

Firstly, there is a significant lack of data collected from working women in rural areas, which undermines the generalizability of the findings across different geographic and socio-economic contexts. Furthermore, the study lacks diversity in the professional backgrounds of the participants, with a predominant focus on teachers. This professional homogeneity limits the applicability of the results to a broader spectrum of occupations, potentially skewing the conclusions drawn from the data. The study could be improved by including a comparison with the parenting styles of non-working women. Consequently, it is essential for future studies to address the limitations by employing larger and more diverse samples, as well as utilizing robust methodologies.

## CONCLUSION

This research concludes the Authoritative parenting style as the most prevalent among working mothers along with various factors where significant association was found with number of children, family income and marital status. Therefore, in order to equip the parents especially mothers with abilities to effectively foster the needs of their children, it is crucial for the stakeholders, policy makers and educationists to develop parenting education programs, counselling sessions and effective recreational facilities to cultivate a supportive and healthy home environment that would ultimately contribute to a better cognitive and mental development of children.

## Data Availability

All data produced in the present study are available upon reasonable request to the authors.

## Notes

### Competing Interest Statement

The authors have declared no competing interest.

### Funding Statement

This study did not receive any funding.

### Author Declarations

Ethical committee of the Department of Community Medicine, Rawalpindi Medical University, Rawalpindi, Pakistan waived ethical approval for this work.

### Summary of Updates

Title updated; Discussion updated; Author affiliations updated

## REFERENCES

1- Howenstein, J., Kumar, A., Casamassimo, P. S., McTigue, D., Coury, D., & Yin, H. (2015). Correlating parenting styles with child behavior and caries. Pediatric Dentistry, 37(1), 59–64

2- Dwairy, M., Achoui, M., Abouserie, R., Farah, A., Sakhleh, A. A., Fayad, M., & Khan, H. K. (2006). Parenting styles in Arab societies: A first cross-regional research study. Journal of Cross-Cultural Psychology, 37(3), 230–247

3- Martínez, I., García, J. F., & Yubero, S. (2007). Parenting styles and adolescents’ self-esteem in Brazil. Psychological Reports, 100(3), 731–745.

4- Hadjicharalambous, D., & Demetriou, L. (2020). The relationship between parents’ demographic factors and parenting styles: Effects on children’s psychological adjustment. Available at SSRN: 10.2139/ssrn.3647329

5- Almudhee, S. N., Al Saigul, A. M., & Sulaiman, A. (2023). Parenting style frequency and their sociodemographic determinants in Buraidah City, Qassim, Saudi Arabia. Cureus, 15(7), e41388. 10.7759/cureus.41388

6- Sanvictores, T., & Mendez, M. D. (2022). Types of parenting styles and effects on children. In StatPearls (2024 ed.). Treasure Island, FL: StatPearls Publishing.

7- Arafat, S. Y., & Khan, M. A. (2022). Parenting style in Bangladesh. Parenting, 4, 5.

8- Huver, R. M. E., Otten, R., De Vries, H., & Engels, R. C. M. E. (2010). Personality and parenting style in parents of adolescents. Journal of Adolescence, 33

9- Krisnana, I., Rachmawati, P. D., Arief, Y. S., Kurnia, I. D., Nastiti, A. A., Safitri, I. F. N., & Putri, A. T. K. (2019). Adolescent characteristics and parenting style as determinant factors of bullying in Indonesia: A cross-sectional study. International Journal of Adolescence and Medicine Health, 33(5). 10.1515/ijamh-2019-0019

